# An online tool for correcting verification bias when validating electronic phenotyping algorithms

**DOI:** 10.1101/2023.11.22.23298913

**Authors:** Ajay Bhasin, Suzette J. Bielinski, Abel N. Kho, Nicholas B. Larson, Laura Rasmussen-Torvik

**Affiliations:** Department of Medicine, Division of Hospital Medicine, Northwestern University Feinberg, School of Medicine, Chicago, IL; Department of Pediatrics, Division of Hospital-Based Medicine, Northwestern University, Feinberg School of Medicine, Chicago, IL; Division of Epidemiology, Department of Quantitative Health Sciences, Mayo Clinic College of Medicine and Science, Rochester, Minnesota, USA; Center for Health Information Partnerships, Institute for Public Health & Medicine, Feinberg, School of Medicine, Northwestern University, Chicago, IL, USA; Division of General Internal Medicine, Department of Medicine, Feinberg School of Medicine, Northwestern University, Chicago, IL, USA; Division of Clinical Trials and Biostatistics, Department of Quantitative Health Sciences, Mayo Clinic College of Medicine and Science, Rochester, MN, USA; Department of Preventive Medicine, Division of Epidemiology, Northwestern University Feinberg School of Medicine

## Abstract

Computable or electronic phenotypes of patient conditions are becoming more commonplace in quality improvement and clinical research. During phenotyping algorithm validation, standard classification performance measures (i.e., sensitivity, specificity, positive predictive value, negative predictive value, and accuracy) are commonly employed. When validation is performed on a randomly sampled patient population, direct estimates of these measures are valid. However, it is common that studies will sample patients conditional on the algorithm result, leading to a form of bias known as verification bias. The presence of verification bias requires adjustment of performance measure estimates to account for this sampling bias. Herein, we describe the appropriate formulae for valid estimates of sensitivity, specificity, and accuracy to account for verification bias. We additionally present an online tool to adjust algorithm performance measures for verification bias by directly taking the sampling strategy into consideration and recommend use of this tool to properly estimate algorithm performance for phenotyping validation studies.

## Introduction

Computable phenotypes of patient conditions are becoming more commonplace in quality improvement and clinical research.^1^ These phenotypes are algorithmically derived from data sources such as electronic health record (EHR), insurance claims, or centers for Medicare and Medicaid Services data, and can empower research and improve patient care.^2,3^ Algorithm performance measures, such as sensitivity, specificity, and positive and negative predictive values (PPV and NPV) are common measures of validity obtained by comparing the algorithm result to a “gold standard” (e.g. manual chart review). A common validation study design strategy when the condition of interest has low prevalence is to sample based on the algorithm result (e.g. 50 predicted cases and 50 predicted non-cases).^4,5^ This strategy is both cost-effective and statistically efficient by enriching for likely true positives and improving the expected precision of positive-class performance measures (e.g., sensitivity, PPV). However, this sampling strategy also results in a form of selection bias known as verification bias, which is commonly encountered in diagnostic test evaluation.^6-8^ Under these conditions, estimates of sensitivity, specificity, and accuracy can be biased if the sampling design is not taken into consideration. Herein, we illustrate the effects of verification bias on performance estimation through an example validation study and develop a user-friendly online tool to facilitate adjustment of performance measures under these validation study scenarios.

## Methods

Given that EHR-based phenotyping algorithms can be prone to error, it is often of interest to characterize classification performance relative to ground truth based on manual chart abstraction. Formulae for defining these performance measures adjusting estimates of sensitivity and specificity for verification bias are available in Figure 1. Detailed explanations of these derivations, along with formulae for calculating corresponding asymptotic CI’s, are provided by Begg and Greenes.^9^

### Validation Study Sampling Design

For phenotyping algorithms, the total number of patients with available classification results tends to be very large due to ease of implementation (e.g., the entire patient population at a medical institution). Given the potential laborious nature of chart review, algorithm validation studies are often performed on a relatively small subset of the total population. When the expected prevalence of the disease condition is low (i.e., less than 10%), validation studies may have correspondingly low precision for estimating sensitivity and PPV if patients are randomly sampled from the population. For example, for a disease with prevalence of 2%, in a random sample of 500 patients we expect 10 positive disease patients, on average. Even at a true algorithm sensitivity of 90% (i.e., 9/10 cases correctly identified), the Wilson score 95% confidence interval (CI) would be [0.596,0.995]. In contrast, 90% specificity would correspond to a 95% confidence interval of [0.870,0.925]. This disparity in precision can be mitigated by oversampling subjects predicted by the algorithm as a positive case (e.g., 1:1 sampling based on predicted disease status), leading to a more balanced representation of true disease cases and unaffected non-cases within the validation sample.

### Naïve and Adjusted Validation Performance

While the sampling strategy defined above leads to more statistically efficient estimation of algorithm performance, sampling patients for the validation study based on algorithm-classified disease status can lead to biased estimation of performance measures. Referred to as “verification” or “work-up” bias, unadjusted analyses of the resulting validation 2x2 contingency table can specifically lead to overestimated sensitivity while simultaneously underestimating specificity. However, since NPV and PPV correspond to probabilities conditional on predicted statuses, these estimates remain valid under this conditional sampling scheme.

### Example Validation Study

Consider the illustrative example of a validation study where a phenotyping algorithm is applied to a source population of 1,100 patients, corresponding to 100 patients classified as positive and 1000 patients as negative. From this cohort, 50 predicted cases and 50 predicted non-cases were selected for phenotyping algorithm validation. The manual abstraction yielded a 2x2 contingency table with counts of 49 true positives, 1 false positive, 3 false negatives, and 47 true negatives.

**Figure 1.**
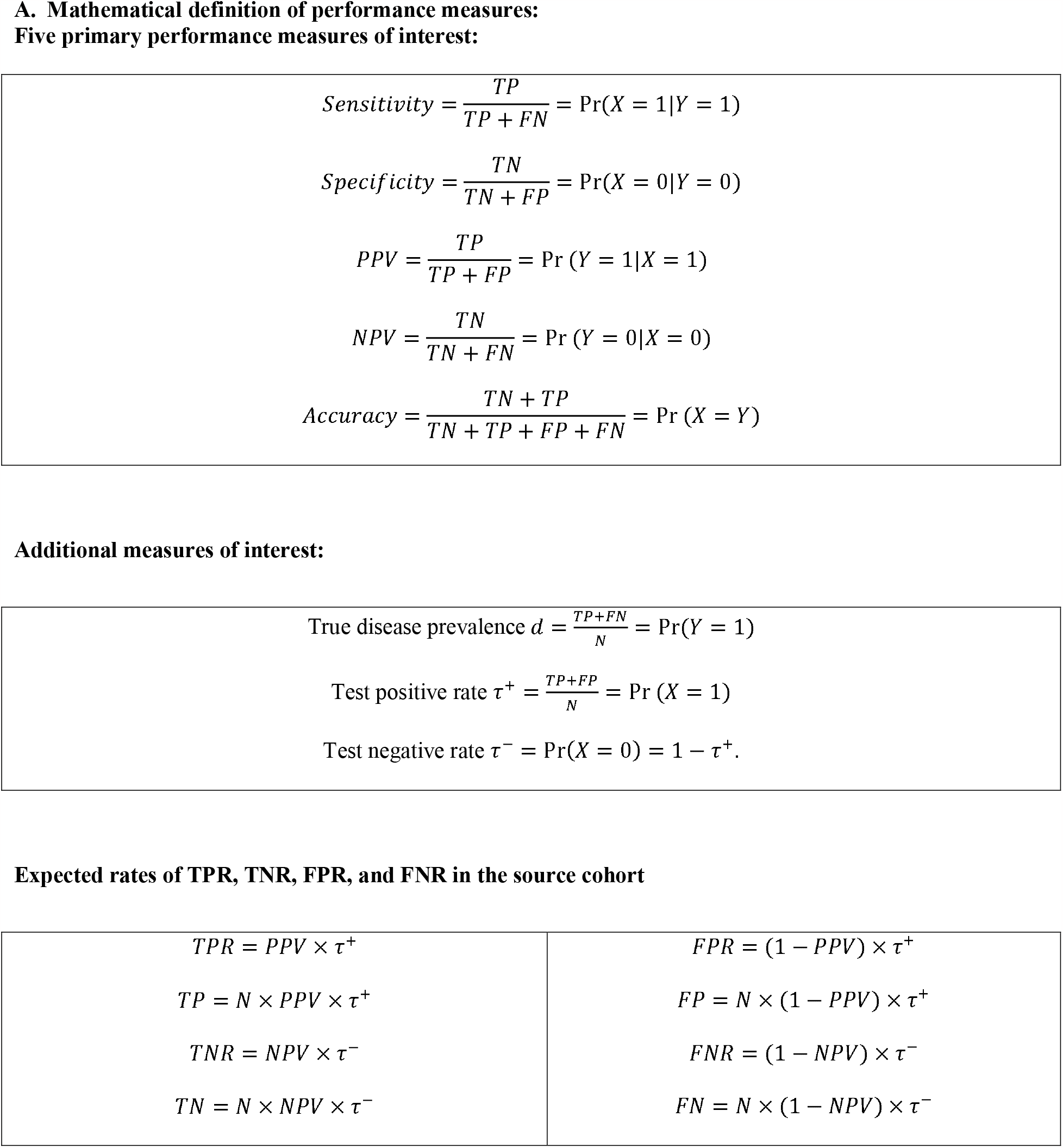
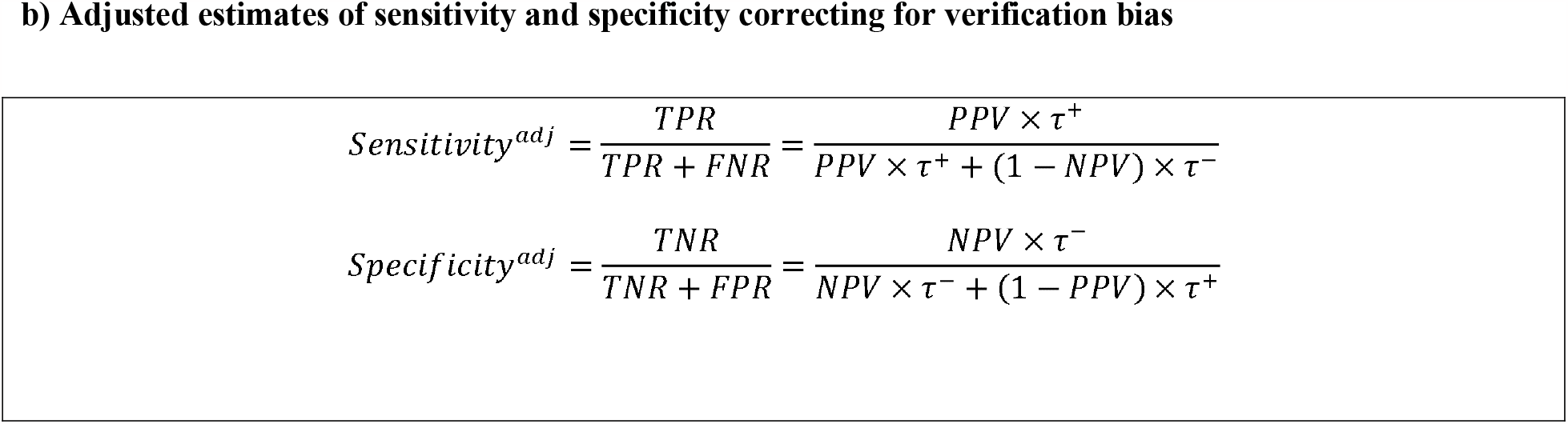
Consider a phenotyping algorithm for predicting the presence of a given disease condition based on a patient’s EHR data. We designate *Y* ∈ {*0,1*} to be the true underlying disease status for a given patient and *X* ∈ {*0,1*} to be the predicted disease status by the algorithm, such that 0 and 1 respectively denote unaffected and affected disease statuses. For disease phenotyping on a patient cohort of size *N*, the classification results can be summarized using a standard 2x2 contingency table, which tabulates patient classifications of disease relative to true disease status into four distinct categories: true positives (TP), true negatives (TN), false positives (FP), and false negatives (FN), as indicated in Table 1. Counts in the equations above can be replaced by corresponding rates by simply factoring out *N* (e.g., the true positive rate 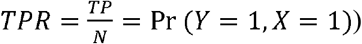. Given that unbiased estimates of test positive and negative rates, *τ*^+^ and *τ*^-^, are available from the algorithm classifications for the original source cohort, the expected rates of TPR, TNR, FPR, and FNR in the source cohort can actually be calculated as simple functions of these parameters and the PPV and NPV estimates from the validation study. For example, recall from above that TPR can be framed as the joint probability Pr(*Y* = 1, *X* = 1). Since Pr(*Y* = 1, *X* = 1) Pr(*Y* = 1|*X* = 1) × Pr(*X* = 1) by basic rules of conditional probability, and Pr(*Y* = 1|*X* = 1) = *PPV* and Pr(*X* = 1) = *τ*^+^per our definitions above, it follows that *TPR* = *PPV* × *τ*^+^

### Simulation Analysis

To further illustrate the impact of verification bias on sensitivity and specificity estimates across a broad range of realistic study conditions, we conducted a simple simulation study for a disease with estimated true prevalence between 1% and 50%; true NPV of 0.90, 0.95, and 0.99; and true PPV of 0.70, 0.80, and 0.90. For validation, we considered a balanced study design, such that equal numbers of predicted cases and non-cases are selected for chart abstraction. We then calculated the expected bias of naive estimates of sensitivity and specificity relative to appropriately adjusted estimates based on expected values of true positive rate (TPR), false positive rate (FPR), true negative rate (TNR), and false negative rate (FNR) in the validation study.

**Table 1:** 2x2 contingency table definitions for phenotyping validation.

|  | Validation Study |  |  | Source Population |  |  |
| --- | --- | --- | --- | --- | --- | --- |
|  | Chart (+) | Chart (-) | Total | Disease | No Disease | Total |
| **Algorithm (+)** | 49 | 1 | 50 | 98 | 2 | 100 |
| **Algorithm (-)** | 3 | 47 | 50 | 60 | 940 | 1000 |
| **Total** | 52 | 48 | 100 | 158 | 942 | 1100 |

### Online Tool

We used Microsoft Visual Studio Code (version 1.78.0) and Python (version 3.10) with the *Streamlit* package (version 1.13.0) to create a simple tool to calculate sensitivity, specificity, PPV, NPV, and accuracy of a phenotyping algorithm based on chart validation. The tool is freely available at: https://bit.ly/3tMTJiE.

## Results

The 2x2 contingency table of the example validation study along with projected counts from the total source cohort are presented in Table 1, while respective performance measure analyses corresponding to unadjusted and verification-bias adjusted estimates are presented in Table 2.

**Table 2:** Comparison of classification performance measures based on unadjusted analysis of the validation study table and verification bias adjusted estimates. Note that PPV and NPV are identical across both analyses.

| Measures | Naïve |  | Bias-Adjusted |  |
| --- | --- | --- | --- | --- |
| **Prevalence** | 0.520 |  | 0.091 |  |
| **Accuracy** | 0.960 |  | 0.944 |  |
| **PPV (95% CI)** | 0.980 | [0.895,0.999] | - | - |
| **NPV (95% CI)** | 0.940 | [0.838,0.979] | - | - |
| **Sensitivity (95% CI)** | 0.942 | [0.844,0.980] | 0.620 | [0.553,0.683] |
| **Specificity (95% CI)** | 0.979 | [0.891,0.999] | 0.998 | [0.997,0.998] |

Unadjusted performance estimates for the hypothesized phenotyping algorithm corresponded to 0.942 sensitivity, 0.979 specificity, and 0.960 accuracy. The disease prevalence in the validation study sample was 0.520, whereas the true prevalence in the source population was 0.091. After adjusting for verification bias, the updated performance measures for the algorithm corresponded to 0.620 sensitivity, 0.999 specificity, and 0.944 accuracy.

Results from our simulation study are presented in Figure 2. These results illustrate the substantial positive bias for sensitivity estimation that may be observed as disease prevalence decreases toward zero when analyzing the unadjusted validation study confusion matrix results. This bias relationship is attenuated as the NPV approaches 1.00, but still yields extreme bias at lower prevalence values. For specificity (Figure 2B), we observe similar trends of increased absolute bias with decreased prevalence. However, the magnitude of this bias remains largely consistent across realistically high values of NPV considered for the simulation study, with lower PPV leading to moderate increases in bias. Of note, these results represented expected biases, and actual results may vary based on sizes of the total population and sampling cohort due to sampling variability.

**Figure 2:**
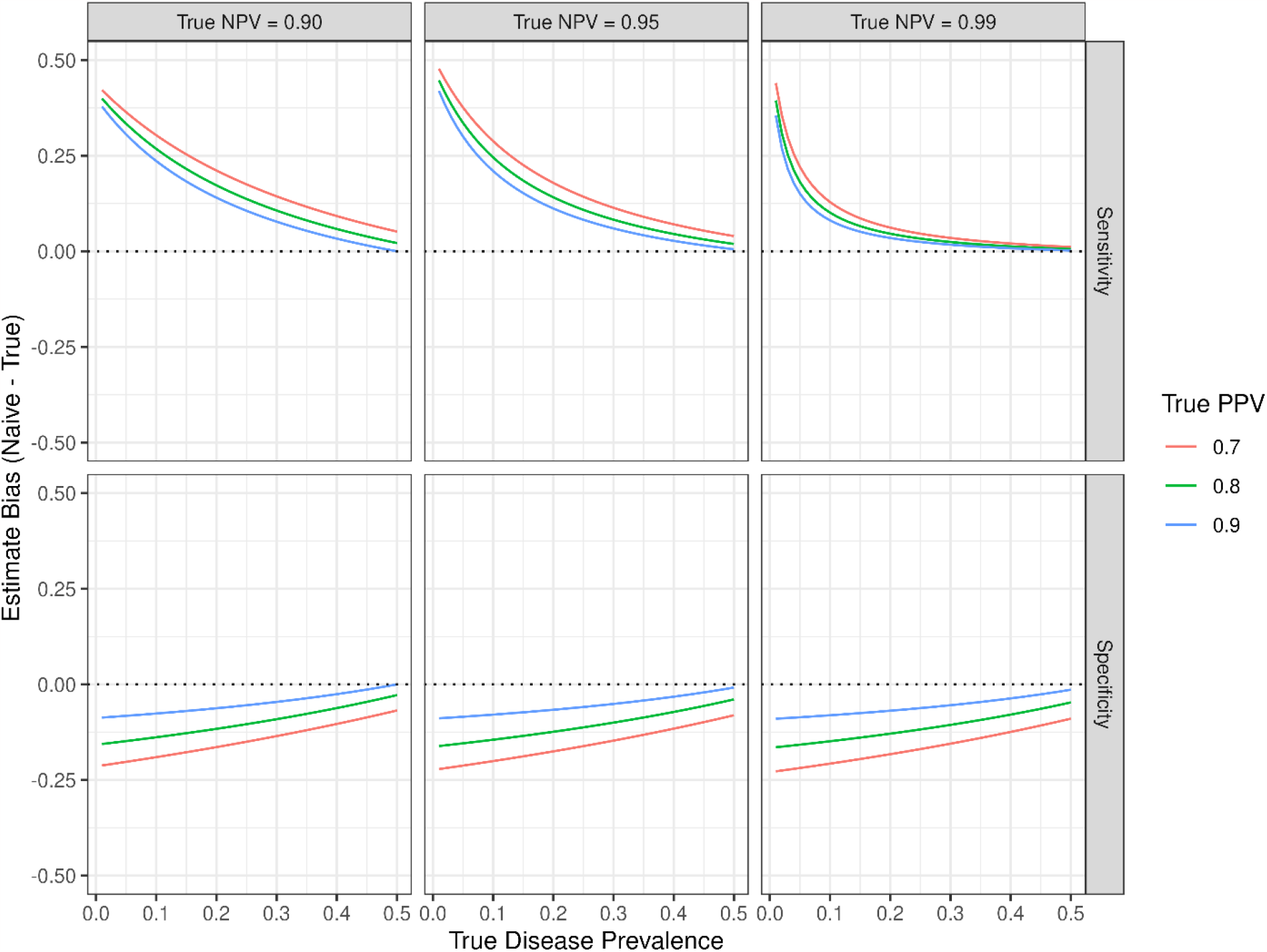
Simulation study results demonstrating expected biases for sensitivity and specificity under verification bias for various values of true PPV, true NPV, and disease prevalence.

## Discussion

The provided example demonstrates the performance metrics of an algorithm and how much they can change when one does not randomly sample from the source population for algorithm validation. Oversampling of algorithm-positive cases for validation can bias model performance measures, leading to inflated sensitivity and accuracy estimates. The bias can be mitigated by considering the prevalence of disease in the source population and adjusting the calculations to account for the difference.

While sampling conditional on predicted disease status will lead to valid direct estimates of PPV and NPV, these measures are themselves a function of disease prevalence. Thus, they are not necessarily intrinsic properties of a phenotyping algorithm, and should be interpreted with caution as disease prevalence may vary across validation populations.^10^ Likewise, alternative performance measures that are in part functions of sensitivity and/or specificity, such as F1-score and positive/negative likelihood ratios, will also likely be biased and require similar corrections. Stratified study designs can also be adopted when there are covariates that may correlate with differential algorithm performance, and we refer the reader to appropriate references for how to address adjustment under these conditions.^6,9^

For accurate adjustment and algorithm calibration, the source population should be defined prior to application of an algorithm. Ideally, a very high percentage of the source population will be characterized by the algorithm: if a high percentage of patients are not classified as either disease positive or negative by the algorithm, then the performance metrics of the algorithm will be difficult to interpret and this will significantly increase the difficulty of cross-institutional validation.^11-13^

This tool will enable clinicians, informaticists, and data scientists to appropriately characterize performance of computable phenotype algorithms.

## Data Availability

There is no data associated with this study. There is an online web tool that we have created that is free to use by anyone reviewing the paper.

https://bit.ly/3tMTJiE

## Notes

**Conflict of Interest Disclosures:** To the best of our knowledge, no conflict of interest, financial or other, exists with respect to the information provided in this report.

**Funding/Support:** None.

### Competing Interest Statement

The authors have declared no competing interest.

### Funding Statement

The study did not receive any funding.

